# Distribution of paramagnetic and diamagnetic cortical substrates following mild Traumatic Brain Injury: A depth- and curvature-based quantitative susceptibility mapping study

**DOI:** 10.1101/2024.08.16.24312140

**Authors:** Christi A. Essex, Jenna L. Merenstein, Devon K. Overson, Trong-Kha Truong, David J. Madden, Mayan J. Bedggood, Helen Murray, Samantha J. Holdsworth, Ashley W. Stewart, Catherine Morgan, Richard L. M. Faull, Patria Hume, Alice Theadom, Mangor Pedersen

## Abstract

Evidence has linked head trauma to increased risk factors for neuropathology, including mechanical deformation of the sulcal fundus and, later, perivascular accumulation of hyperphosphorylated tau adjacent to these spaces related to chronic traumatic encephalopathy (CTE). However, little is known about microstructural abnormalities and cellular dyshomeostasis at the acute stage of mild traumatic brain injury (mTBI) in humans, particularly in the cortex. To address this gap, we designed the first architectonically motivated quantitative susceptibility mapping (QSM) study to assess regional patterns of positive (iron-related) and negative (myelin-, calcium-, and protein-related) magnetic susceptibility across 34 cortical regions of interest following mTBI. Bilateral, between-group analyses sensitive to cortical depth and curvature were conducted between 25 males with acute (*<* 14 days) sports-related mTBI (sr-mTBI) and 25 age-matched male controls. Results suggest a trauma-induced increase in positive susceptibility focal to superficial, perivascular-adjacent spaces in the parahippocampal sulcus. Co-localised decreases in diamagnetism indicate a dual pathology of neural substrates. These mTBI-related patterns were distinct from age-related processes, as revealed by correlation analyses. Our findings suggest depth- and curvature-specific deposition of biological substrates in cortical tissue following mTBI, and the coherence between the superficial sulcal predilection and pathognomonic patterns of misfolded proteins in trauma-related neurodegeneration is interesting.

## Introduction

Mild traumatic brain injury (mTBI) is responsible for up to 90% of the estimated 27-69 million annual incidents of traumatic brain injury worldwide (Dewan et al., 2019, James et al., 2019, Maas et al., 2022); representing a *∼*USD $400 billion global economic burden (Howe et al., 2022). In addition to economic impacts, exposure to mild head trauma is a major public health concern. mTBI is associated with adverse mental health effects, cognitive decline, increased risk of neurodegenerative disease (Guskiewicz et al., 2007, McInnes et al., 2017), and premature mortality (Mackay et al., 2019). Repeated instances of brain trauma is a well documented risk factor for the progressive tauopathy known as chronic traumatic encephalopathy (CTE), most often observed in athletes as a result of participation in contact sports, or in military veterans from exposure to blast impacts (Bieniek et al., 2020, 2015, Ling et al., 2017, McKee et al., 2013, 2023, Mez et al., 2017, 2020). mTBI is characterised by a primary insult to the brain, affecting tissue microstructure and inducing a cascade of secondary cellular processes, transient states of metabolic distress, and cellular dyshomeostasis (Giza and Hovda, 2014).

The absence of discernible focal lesions or other macroscopic morphological abnormalities means standard neuroimaging approaches are often insufficiently sensitive to detect mTBI pathology. Advanced neuroimaging methods extending beyond the scope of conventional medical practices may be used to identify the subtle, diffuse (Hier et al., 2021, Lunkova et al., 2021), and heterogeneous changes in brain structure (Cook and Hawley, 2014, Wintermark et al., 2015). Despite promising advances toward discovery of mTBI biomarkers, to date no specific markers of structural brain injury have been identified and assessments are thus restricted to clinical evaluations and self-reported impairments in cognitive and physiological function (Lunkova et al., 2021, McCrea et al., 2017). These assessments may not accurately reflect objective measures of brain injury or recovery status, which can constrain intervention efficacy (McCrea et al., 2017, Shenton et al., 2012).

Investigating the role of biomaterials involved in preserving neuronal homeostasis, including trace elements such as brain iron, represents a promising direction for biomarker research. Iron is essential for proper brain function, and is a cofactor in various neuronal processes including neurotransmitter-, myelin-, and DNA-synthesis, energy metabolism, and oxidative phosphorylation (Ward et al., 2014b). Cellular concentrations are tightly regulated within neural tissue and dyshomeostasis has been linked to oxidative stress, DNA and protein damage, inflammation, and ferroptosis (a form of iron-regulated cell death) (Ma et al., 2022, Mackenzie et al., 2008, Nisenbaum et al., 2014). Evidence suggests iron overload is implicated in both secondary injury and the emergence of pathology downstream (Gozt et al., 2021) including hyperphosphorylation of tau (p-tau) (Yamamoto et al., 2002), highlighting the direct interplay between elevated levels of iron and CTE-like processes. Histologically-validated co-localisation of iron with abnormal protein accumulations in progressive tauopathies has distinguished brain iron as a hallmark feature of degenerative disorders such as Alzheimer’s and Parkinson’s disease (AD and PD, respectively) (Stankiewicz et al., 2007, Zecca et al., 2004) and has been cited within neurofibrillary tangles in CTE (Bouras et al., 1997). Despite evidence linking head trauma to increased risk factors for premature iron-related neuropathology (Daglas and Adlard, 2018) and increased levels of brain iron in humans following mTBI (Nisenbaum et al., 2014), the association between brain iron accumulation and the pathophysiology of acute sports-related mild traumatic brain injury (sr-mTBI) remains unclear.

Quantitative susceptibility mapping (QSM), an advanced magnetic resonance imaging (MRI) method, measures intrinsic magnetic properties and spatial distributions of biomaterials and molecules (including iron, calcium, protein, and myelin) that are related to brain tissue composition (Duyn and Schenck, 2017, Jang et al., 2021, Kim et al., 2020, Wang et al., 2017, Gong et al., 2019, Zhao et al., 2021). The magnetic susceptibility of these materials is proportional to the degree of magnetisation exhibited in response to an external magnetic field, such as the main magnetic field of an MRI scanner (B_0_) (Schweser et al., 2016). Differences in magnetic field perturbation from dia- and para-magnetic compounds in brain tissue create inhomogeneities in the phase maps of gradient-recalled echo (GRE) MRI sequences (De Rochefort et al., 2010, Langkammer et al., 2015, Marques and Bowtell, 2005). This mechanism generates contrast in QSM, thereby providing insights into the architecture of the brain and enabling more accurate delineation of many structural boundaries than the corresponding GRE magnitude images (Liu et al., 2015b).

Unlike traditional susceptibility-weighted imaging (SWI) from which QSM is derived, this approach can be used to directly quantify the susceptibility of tissue within regions of interest (ROIs), serving as a close approximation of the constituent elements (Deistung et al., 2017, Liu et al., 2015a). Already integral to dementia research (Ghaderi et al., 2024, Mohammadi et al., 2024, Nikparast et al., 2022, Ravanfar et al., 2021, Suresh Paul et al., 2024, Uchida et al., 2022), QSM can be extended to investigate potential susceptibility-related pathology resulting from acute sr-mTBI.

A limited number of studies have used QSM to quantify magnetic susceptibility in white matter and/or subcortical or global grey matter (Brett et al., 2021, Gong et al., 2018, Koch et al., 2018, 2021, Weber et al., 2018, Wright et al., 2022, Pinky et al., 2022), or as a marker of cerebral venous oxygen saturation (Sv0_2_) (Chai et al., 2017, Wright et al., 2022, To et al., 2024). QSM investigations of mTBI-related grey matter alterations have focussed almost exclusively on deep grey nuclei as a proxy for injury effects and cellular degeneration. This is due to the high density of iron in these nuclei related to elevated metabolic demand (Gozt et al., 2021), particularly in the globus pallidus, red nucleus, substantia nigra, putamen, dentate, caudate, and thalamus, relative to the cortical grey matter (Hallgren and Sourander, 1958). These deep grey matter sites are not only vulnerable to iron-mediated disorders (Haacke et al., 2005) but also damage in mTBI (Raz et al., 2011). However, studies using QSM to investigate mTBI effects have largely overlooked the cortex. This complex structure is characterised by ridges (gyri) and grooves (sulci); curvatures corresponding to the base (fundus) of these sulci are exposed to the greatest force during mTBI, a phenomenon referred to as the ‘water hammer effect’(Kornguth et al., 2017).

These cortical, microvascular-adjacent, regions most susceptible to mechanical deformation and injury in mTBI are also the primary loci of degeneration and tauopathy in CTE (McKee et al., 2023, Smith et al., 2013) and warrant careful investigation. The singular mTBI-QSM study to include cortical grey matter ROIs was constrained by a macroscopic voxel-wise approach (Gong et al., 2018). Significantly more anatomical precision is needed to detect depth-or curvature-specific differences in magnetic susceptibility within the cortical mantle after mTBI. The cortex is likely excluded from investigation due to several methodological challenges primarily related to complex cortical architectonics. Firstly, differentiating myeloarchitecture, cytoarchitecture, and cortical laminae using ultra-high field, ultra-high resolution MRI (such as that acquired at 7T or higher), is a developing area in neuroimaging research (Waehnert et al., 2016). Currently, however, most research institutions primarily use MRI scanners with lower field strengths. Secondly, standard voxel-wise comparisons are naïve to the architectonics and distribution of cellular elements within the cortical layers; here, advanced analytic techniques are essential. At magnetic field strengths of 3T or lower, and supra-millimetre voxel resolutions, analysis of specific cortical laminae is inhibited; however, column-based analytic techniques (Ma et al., 2023, Merenstein et al., 2024, Northall et al., 2023, Waehnert et al., 2014) enable depth-wise investigations of magnetic susceptibility in the cerebral cortex and are already producing promising results in Alzheimer’s research (Merenstein et al., 2024).

To address these research gaps, we conducted the first architectonically-motivated QSM analysis of cerebral mTBI effects. This study aimed to: 1) assess regional patterns of positive (iron-related) and negative (myelin-, protein-, calcium-related) magnetic susceptibility as a marker of acute cortical pathology after sr-mTBI, and; 2) understand the relationship between magnetic susceptibility in the cerebral cortex and variables such as age, injury latency, and severity. Based on prior literature, we hypothesised that differences in susceptibility would likely be evident in the frontal and temporal cortices, which are reported to be susceptible to injury in mTBI and are among the first to show degenerative effects of brain injury. We expected this distribution to be most prominent in the sulcal fundus due to increased vulnerability to trauma-induced deformation. Based on known effects of age on cortical iron deposition in this age range (Hallgren and Sourander, 1958), positive susceptibility values were hypothesised to show a positive relationship with age; however, correlation analyses remained largely exploratory and without specific *a priori* hypotheses.

## Materials and Methods

Approval for this study was granted by both The Auckland University of Technology Ethics Committee ((AUTEC) Date: 18/02/2022, Ref: 22/12) and the Health and Disabilities Ethics Committee ((HDEC) Date: 18/02/2022, Ref: 2022 EXP 11078). The study was conducted following the Declaration of Helsinki and all participants provided written informed consent prior to data collection. All participants were provisioned a $50NZD food voucher to acknowledge their contribution as well as a $20NZD fuel voucher or taxi chit to cover travel expenses related to MRI scan attendance.

### Participants

Data from 25 male contact sports players (*M* = 21.10 years old [16-32], *SD* = 4.43) with acute sr-mTBI (*<* 14 days; *M* = 10.40 days, *SD* = 3.03) and 25 age-matched male controls (*M* = 21.10 years old [16-32], *SD* = 4.35) were used for this observational, case-control study (see *Table 1*). Clinical (sr-mTBI) participants were recruited through three Axis Sports Medicine clinics (Auckland, New Zealand) and through community-based pathways including referrals from healthcare professionals and sports team management. All clinical participants received a confirmed diagnosis of sr-mTBI by a licensed physician as a prerequisite for study inclusion and symptom severity was assessed with the Brain Injury Screening Tool (BIST) (Theadom et al., 2021) upon presentation to Axis clinics or electronically after recruitment. Healthy controls were recruited through print and social media advertisements, and word-of-mouth. A history of significant medical or neurological conditions unrelated to the scope of this study or contraindication for MRI precluded study participation. Additionally, controls were excluded if they had any recent history of mTBI events (*<* 12 months) or were living with any long-term effects of previous mTBI. All participants completed a 1 hour MRI scan and a short demographic questionnaire. All MRI testing was conducted at The Centre for Advanced MRI (CAMRI), Auckland, New Zealand and relevant scans were reviewed for clinically significant findings by a certified radiologist to ensure participant safety.

**Table 1.**
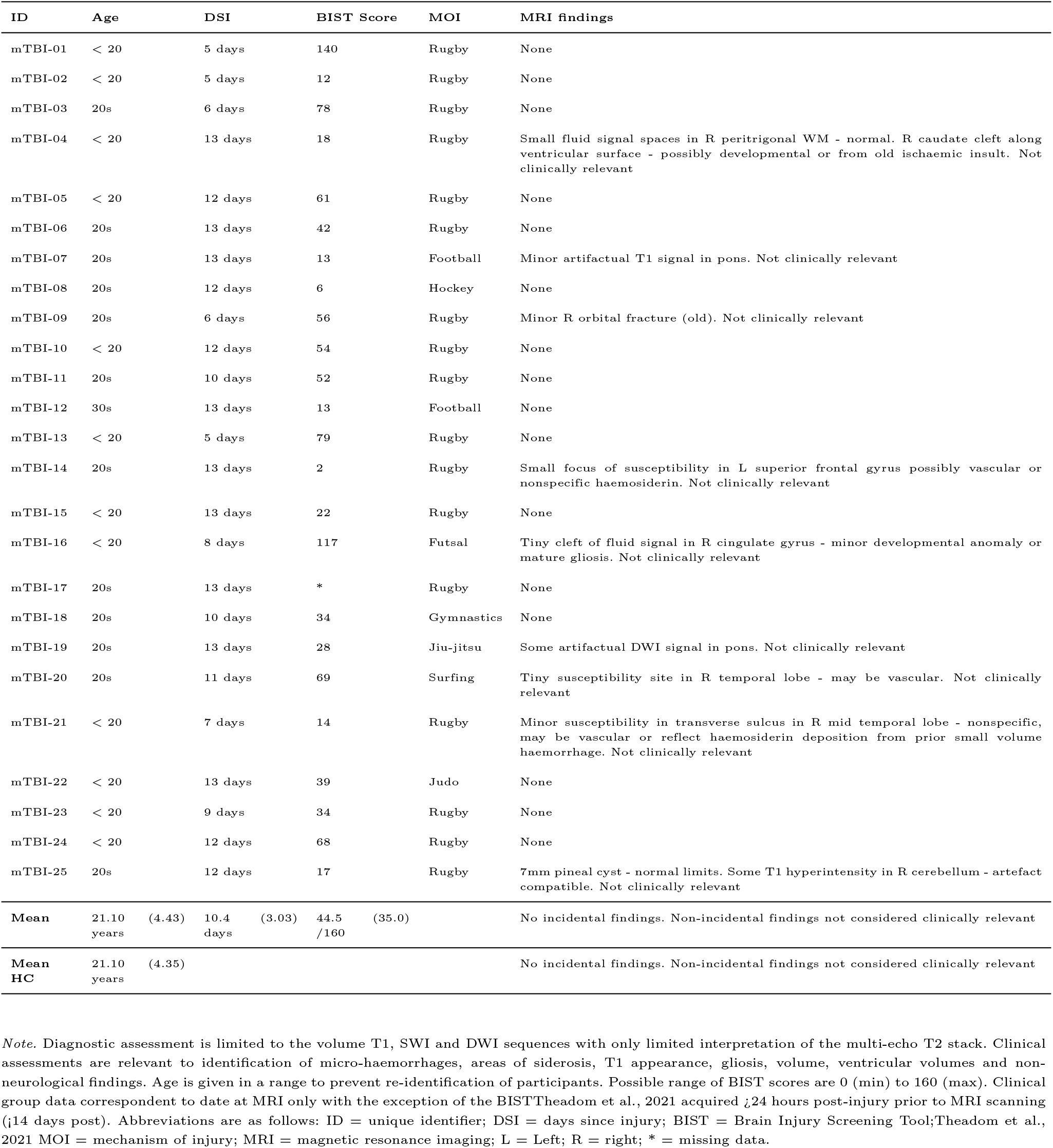
Summary of sr-mTBI participant clinical characteristics with reference to healthy controls.

| ID | Age | DSI | BIST Score | MOI | MRI findings |
| --- | --- | --- | --- | --- | --- |
| mTBI-01 | < 20 | 5 days | 140 | Rugby | None |
| mTBI-02 | < 20 | 5 days | 12 | Rugby | None |
| mTBI-03 | 20s | 6 days | 78 | Rugby | None |
| mTBI-04 | < 20 | 13 days | 18 | Rugby | Small fluid signal spaces in R peritrigonal WM - normal. R caudate cleft along ventricular surface - possibly developmental or from old ischaemic insult. Not clinically relevant |
| mTBI-05 | < 20 | 12 days | 61 | Rugby | None |
| mTBI-06 | 20s | 13 days | 42 | Rugby | None |
| mTBI-07 | 20s | 13 days | 13 | Football | Minor artifactual T1 signal in pons. Not clinically relevant |
| mTBI-08 | 20s | 12 days | 6 | Hockey | None |
| mTBI-09 | 20s | 6 days | 56 | Rugby | Minor R orbital fracture (old). Not clinically relevant |
| mTBI-10 | < 20 | 12 days | 54 | Rugby | None |
| mTBI-11 | 20s | 10 days | 52 | Rugby | None |
| mTBI-12 | 30s | 13 days | 13 | Football | None |
| mTBI-13 | < 20 | 5 days | 79 | Rugby | None |
| mTBI-14 | 20s | 13 days | 2 | Rugby | Small focus of susceptibility in L superior frontal gyrus possibly vascular or nonspecific haemosiderin. Not clinically relevant |
| mTBI-15 | < 20 | 13 days | 22 | Rugby | None |
| mTBI-16 | < 20 | 8 days | 117 | Futsal | Tiny cleft of fluid signal in R cingulate gyrus - minor developmental anomaly or mature gliosis. Not clinically relevant |
| mTBI-17 | 20s | 13 days | * | Rugby | None |
| mTBI-18 | 20s | 10 days | 34 | Gymnastics | None |
| mTBI-19 | 20s | 13 days | 28 | Jiu-jitsu | Some artifactual DWI signal in pons. Not clinically relevant |
| mTBI-20 | 20s | 11 days | 69 | Surfing | Tiny susceptibility site in R temporal lobe - may be vascular. Not clinically relevant |
| mTBI-21 | < 20 | 7 days | 14 | Rugby | Minor susceptibility in transverse sulcus in R mid temporal lobe - nonspecific, may be vascular or reflect haemosiderin deposition from prior small volume haemorrhage. Not clinically relevant |
| mTBI-22 | < 20 | 13 days | 39 | Judo | None |
| mTBI-23 | < 20 | 9 days | 34 | Rugby | None |
| mTBI-24 | < 20 | 12 days | 68 | Rugby | None |
| mTBI-25 | 20s | 12 days | 17 | Rugby | 7mm pineal cyst - normal limits. Some T1 hyperintensity in R cerebellum - artefact compatible. Not clinically relevant |
| **Mean** | 21.10 years (4.43) | 10.4 days (3.03) | 44.5 /160 (35.0) |  | No incidental findings. Non-incidental findings not considered clinically relevant |
| **Mean HC** | 21.10 years (4.35) |  |  |  | No incidental findings. Non-incidental findings not considered clinically relevant |

*Note.* Diagnostic assessment is limited to the volume T1, SWI and DWI sequences with only limited interpretation of the multi-echo T2 stack. Clinical assessments are relevant to identification of micro-haemorrhages, areas of siderosis, T1 appearance, gliosis, volume, ventricular volumes and non-neurological findings. Age is given in a range to prevent re-identification of participants. Possible range of BIST scores are 0 (min) to 160 (max). Clinical group data correspondent to date at MRI only with the exception of the BISTTheadom et al., 2021 acquired <24 hours post-injury prior to MRI scanning (<14 days post). Abbreviations are as follows: ID = unique identifier; DSI = days since injury; BIST = Brain Injury Screening Tool;Theadom et al., 2021 MOI = mechanism of injury; MRI = magnetic resonance imaging; L = Left; R = right; \* = missing data.

### Neuroimaging

#### Acquisition

Advanced MRI data were acquired on a 3T Siemens MAGNETOM Vida Fit scanner (Siemens Healthcare, Erlangen, Germany) equipped with a 20-channel head coil. A 3D flow-compensated GRE sequence was used to obtain magnitude and phase images for QSM reconstruction at 1 mm isotropic resolution (TR = 30 ms; TE = 20 ms; FA = 15^*°*^; slice thickness = 1.0 mm; FoV = 224 mm; matrix size = 180 × 224 × 160 mm) for a total acquisition time of *∼* 3.43 minutes. For each participant, a high-resolution 3D T1-weighted (T1w) anatomical image volume was acquired for coregistration, parcellation and segmentation using a Magnetisation-Prepared Rapid Acquisition Gradient Echo (MPRAGE) sequence (TR = 1940.0 ms; TE = 2.49 ms, FA= 9^*°*^; slice thickness = 9 mm; FoV = 230 mm; matrix size = 192 × 512 × 512 mm; GRAPPA = 2; voxel size 0.45 × 0.45 × 0.90 mm) for a total acquisition time of *∼* 4.31 minutes. T2 maps, resting-state functional MRI data, diffusion-weighted images, and susceptibility-weighted images were also acquired as part of a larger study and analysed separately. Participant DICOM images were converted to NIfTI files and transformed to brain imaging data structure (BIDS) (Gorgolewski et al., 2016) for further processing using *Dcm2Bids* (Boré et al., 2023) version 3.1.1, which is a wrapper for *dcm2niix* (Li et al., 2016) (v1.0.20230411).

#### Anatomical image processing

First, bias field correction was performed on the T1w images for each subject using the *N4* algorithm (Tustison et al., 2010) from ANTs (Avants et al., 2011). The bias field-corrected T1w images were processed in FreeSurfer (Fischl, 2012) to 1) delineate pial and grey matter/white matter (GM/WM) boundary meshes, and; 2) generate estimates of cortical thickness and curvature for each vertex (Merenstein et al., 2024). Skull-stripping was re-run with additional arguments, including *-gcut* and adjustments to the watershed threshold as needed, to improve the accuracy of the original FreeSurfer (Fischl, 2012) pial surface mesh. The pipeline for T1w and QSM image processing is summarised in *Fig. 1*.

**Fig. 1.**
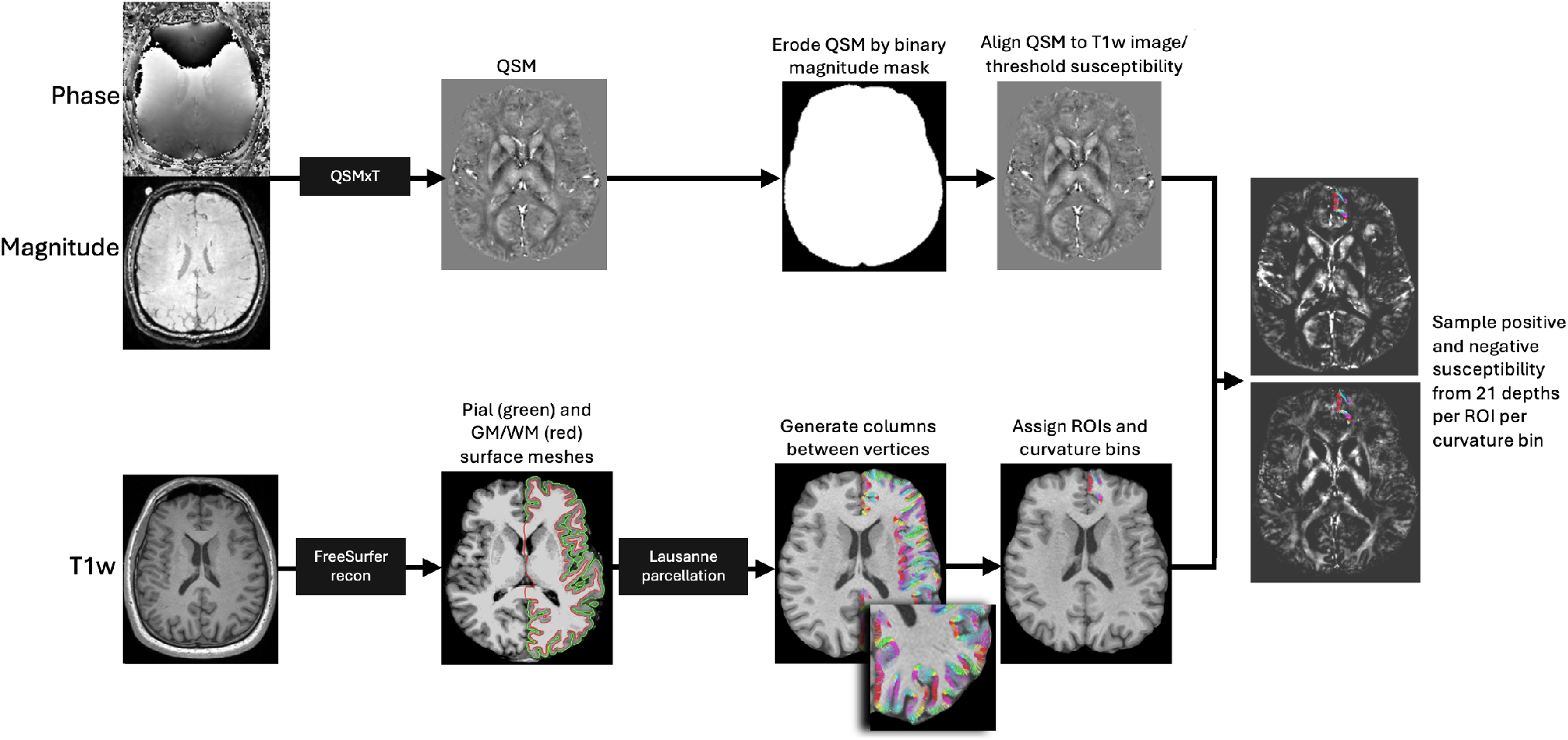
Image processing pipeline. Steps are performed independently and in parallel for each participant. Quantitative susceptibility maps were reconstructed from magnitude and phase images using QSMxT, and eroded by the skull-stripped, binarised magnitude image mask to remove non-brain sources of susceptibility. QSM images were then aligned to T1w images, and thresholded into positive and negative susceptibility maps. FreeSurfer recon was used on bias field-corrected T1w images to produce pial and GM/WM boundary surface meshes, and vertex pairs were then joined to create cortical columns. Parcellations of the cortical grey matter were estimated by feeding the T1w FreeSurfer recon into *easy lausanne*. Columns were then assigned specific ROIs and curvature bins, and used to sample susceptibility from thresholded QSM maps.

#### QSM processing

QSM images were reconstructed using QSMxT (Stewart and Bollman, 2022) v6.4.2 (https://qsmxt.github.io/QSMxT/) available as a container via NeuroDesk (Renton et al., 2024) (v2024-03-27), a lightweight virtual environment. QSMxT integrates and automates phase unwrapping using a rapid open-source minimum spanning tree algorithm (ROMEO) (Dymerska et al., 2021), background field removal with projection onto dipole fields (PDF) (Liu et al., 2011), and sparsity-based rapid two-step dipole inversion (RTS) (Kames et al., 2018); a pipeline congruent with recent consensus statement recommendations for best-practice QSM reconstruction (Bilgic et al., 2023). QSMxT also enables a novel two-pass combination method for hole-filling and artefact reduction (Stewart et al., 2022) which performs parallel QSM masking and reconstruction on susceptibility sources identified as reliable and less reliable. Dual QSM images are then combined into a final integrated image more robust to reconstruction errors and streaking artefacts than those produced using a single-pass approach (Stewart et al., 2022). A brain mask was also generated using FSL’s *BET* (Smith, 2002) to improve masking and hole-filling of the threshold-based selection algorithm (Otsu, 1979) used for two-pass QSM.

Subsequent processing was performed locally using FSL (Jenkinson et al., 2012, Smith et al., 2004, Woolrich et al., 2009). For each subject, the raw magnitude image was skull-stripped using FSL’s *BET* (Smith, 2002) with robust brain centre estimation and a fractional intensity threshold of between 0.3 and 0.6. Binary masks were derived from the skull-stripped magnitude image and applied to the susceptibility maps to erode non-brain noise around the brain perimeter using *fslmaths*. Skull-stripped T1w images were used for the linear coregistration of the magnitude image using FMRIB’s Linear Image Registration Tool (*FLIRT*) (Greve and Fischl, 2009, Jenkinson et al., 2002, Jenkinson and Smith, 2001) with 12 degrees of freedom (DoF). Due to variability in acquisition parameters, FoV, and matrix size between subject images, the 12 DoF linear registration provided more accurate alignment compared to the 6 DoF alternative, allowing for better compensation of non-rigid anatomical variations upon visual inspection. The resulting transformation matrix was used for spatial normalisation of the QSM images to T1w space, effectively upsampling the QSM images. As the analyses are based on cortical depth rather than voxel-wise comparisons, upsampling was not considered a concern. In line with prior research (Merenstein et al., 2024), QSM maps were then thresholded into separate sign (positive and negative) susceptibility maps with *fslmaths*. Traditional QSM maps represent average voxel-wise susceptibility distributions (Reichenbach, 2012), thereby obscuring individual susceptibility sources. Thresholding may enable better distinction of materials with varying biomagnetic properties (e.g., paramagnetic (likely iron) and diamagnetic (likely myelin, calcium, and protein)), and may facilitate a more precise estimation of neurobiological characteristics in analyses.

#### Cortical column generation

To generate cortical columns and sample signed susceptibility values, we used a pipeline previously applied to DWI data analysis (Ma et al., 2023) and recently adapted for use with QSM (Merenstein et al., 2024). First, the T1w FreeSurfer (Fischl, 2012) recon served as an input into the *easy lausanne* tool (https://github.com/mattcieslak/easy lausanne.git). This stripped-down fork of the open-source Connectome Mapper (Daducci et al., 2012) separates the cortex into five atlases, ranging from 34 to 250 ROIs per hemisphere, according to the Lausanne multi-scale atlas (Cammoun et al., 2012). For subsequent analyses, we focused on the atlas with 34 ROIs per hemisphere, which is equivalent to the Desikan-Killiany atlas (Desikan et al., 2006) native to FreeSurfer (Fischl, 2012).

Cortical columns were created for each hemisphere in T1w space with *write mrtrix tracks* (Tournier et al., *2019)* in MATLAB (version R2024a), which was used to connect vertex pairs between the pial and GM/WM boundary surface meshes. Each cortical column was segmented into 21 equidistant depths, each with a step size of 5% of the cortical thickness (Waehnert et al., 2014, 2016), from the pial surface to the GM/WM boundary using MRtrix3 tckresample (Tournier et al., 2019). Here, the step-wise depths represent the 21 equidistant segmentations rather than specific cellular laminae (L1 to L6) of the cortex. It is important to distinguish results produced using this approach from ultra-high field investigations of cyto- and myelo-architecture in the cerebral cortex; results described herein are related to cortical depth, rather than *layer*. The columns were categorised based on cortical curvature, derived from FreeSurfer’s (Fischl, 2012) Gaussian curvature values at each GM/WM boundary vertex (Pienaar et al., 2008) and quantified in units of 1/mm^2^. The categories included the gyral crown (curvature values: 0.6 to 0.1), sulcal bank (0.1 to 0.1), and sulcal fundus (0.1 to 0.6) (Merenstein et al., 2024). Positive curvature values indicated sulci, while negative values indicated gyri, with higher values corresponding to deeper curvatures (Merenstein et al., 2024). Only columns ranging from 0.5 mm to 6 mm in length were included in the analysis to capture plausible cortical morphology (Fischl et al., 2000). Depth was measured in percentage of cortical thickness rather than absolute metrics (mm) to mitigate any variability between control and clinical participants.

### Statistical analyses

To provide a detailed analysis of microstructural differences associated with sr-mTBI while maintaining result granularity, we performed analyses at the bilateral regional level using MATLAB (2024a). Average positive and negative susceptibility values were extracted from 21 cortical depths for all 34 ROIs. Each ROI was analysed by independent curvature bin (gyral crown, sulcal bank, and sulcal fundus) as well as combined curvature as a whole-ROI measure. To control for multiple comparisons and align with prior research (Merenstein et al., 2024) we applied a false discovery rate (FDR) correction (Benjamini and Hochberg, 1995) to the p-values for 21 comparisons (one for each depth) for each ROI/curvature profile. Due to precise age-matching of participants, age was not considered a covariate or confounding variable of interest for between-group comparisons. However, to explore the relationship between QSM values and age in the entire sample, partial Pearson correlation coefficients were calculated between age and both positive and negative susceptibility values independently for all 34 curvature-combined ROIs, at each depth, whilst controlling for group effects. To explore the relationship between susceptibility values and other sr-mTBI-related variables, Pearson correlation coefficients were also calculated between Brain Injury Screening Tool (BIST) (Theadom et al., 2021) scores and injury latency (days since injury (DSI)) and both positive and negative susceptibility values independently for the the sr-mTBI sample only. mTBI-17 was excluded from correlations between BIST and both susceptibility signs due to missing data. Correlations were also corrected for 21 depth-wise comparisons using FDR procedures (Benjamini and Hochberg, 1995). Given the limited sample size and the need to conserve degrees of freedom in this exploratory study, regression analyses were deliberately omitted.

## Results

### Regional depth- and curvature-based analyses

Using independent sample t-tests, we examined depth- and curvature-specific between group differences in bilateral regional susceptibility (positive and negative) for each of the 34 ROIs (see *Fig. 2*). P-values were corrected for 21 cortical depths for each ROI/curvature combination using FDR procedures (Benjamini and Hochberg, 1995).

**Fig. 2.**
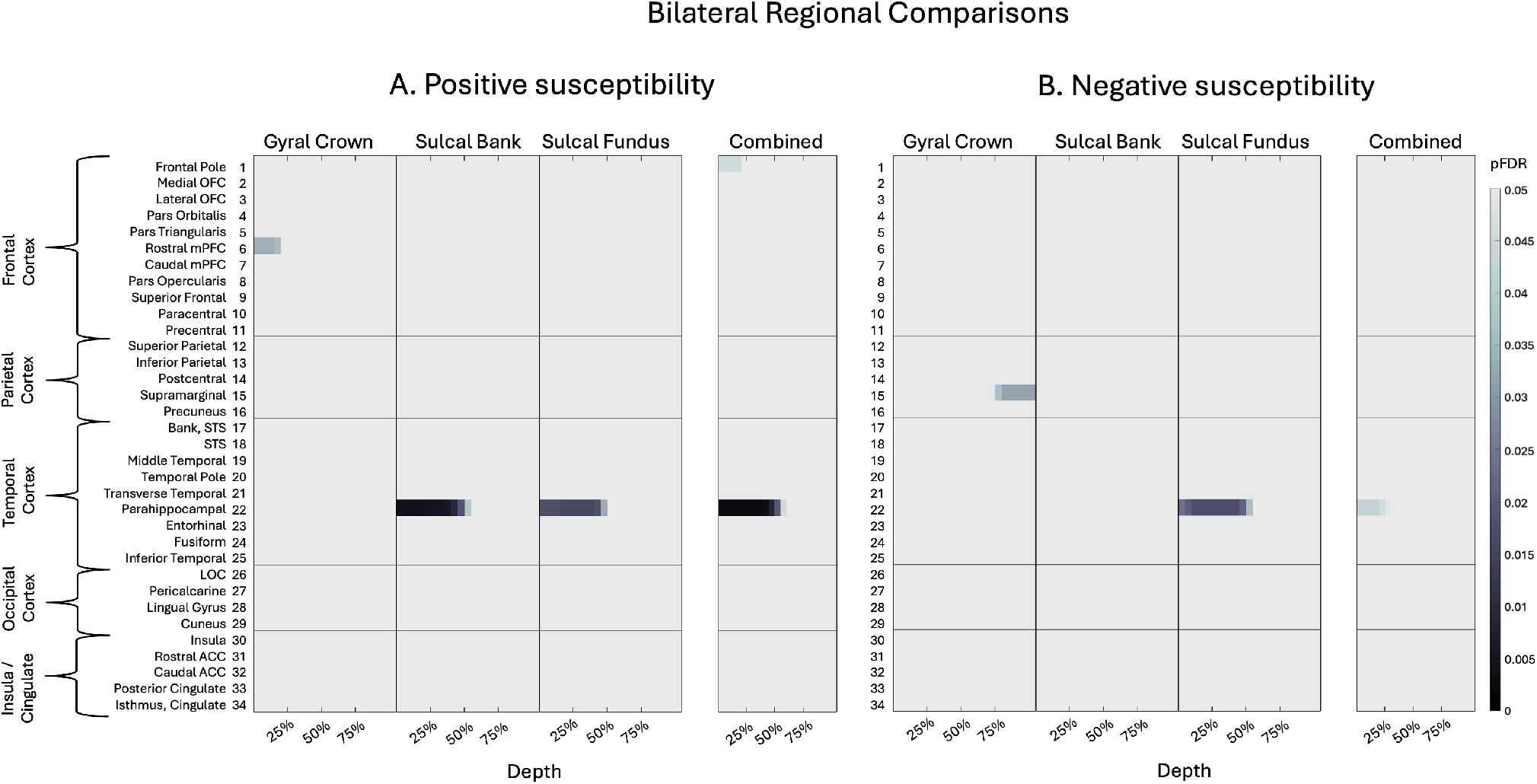
Bilateral regional curvature and depth results. Colour maps demonstrate differences in magnetic susceptibility at each cortical depth (where 0% depth is proximal to the pial surface and increases toward the GM/WM boundary at 100% depth), separately for each curvature bin (crown, bank, fundus) as well as combined curvature as a whole-ROI measure. Independent sample t-tests examined group differences in susceptibility at each depth, for each curvature, at each ROI. P-values were corrected for multiple comparisons across 21 cortical depths using FDR. A. Positive susceptibility values were significantly more positive for participants with sr-mTBI than healthy controls in temporal ROIs and significantly less positive in frontal regions only. B. Susceptibility values were significantly less negative for sr-mTBI participants than controls in temporal ROIs and significantly more negative in parietal ROIs. OFC = orbitofrontal cortex; mPFC = middle prefrontal cortex; STS = superior temporal sulcus; LOC = lateral occipital cortex; ACC = anterior cingulate cortex. Figure based on Merenstein et al. (2024). Depth- and curvature-based quantitative susceptibility mapping analyses of cortical iron in Alzheimer’s disease. *Cerebral Cortex*.

#### Positive susceptibility

Across bilateral depth profiles, participants with sr-mTBI exhibited significantly higher positive susceptibility than controls in the temporal lobe only (see *Fig. 2A*), specifically in superficial depths of the sulcal bank and fundus of the parahippocampal gyrus, a finding that was conserved when curvature was combined as a whole-ROI measure (see also *Fig. 3*.*1A* and *Fig. 3*.*2A*). Susceptibility was decreased following sr-mTBI in the superficial gyral crown of the rostral medial prefrontal cortex (mPFC) and superficially in the frontal pole when curvature was combined. No significant differences between groups were found in bilateral susceptibility values in parietal, occipital, or insular lobes.

**Fig. 3.**
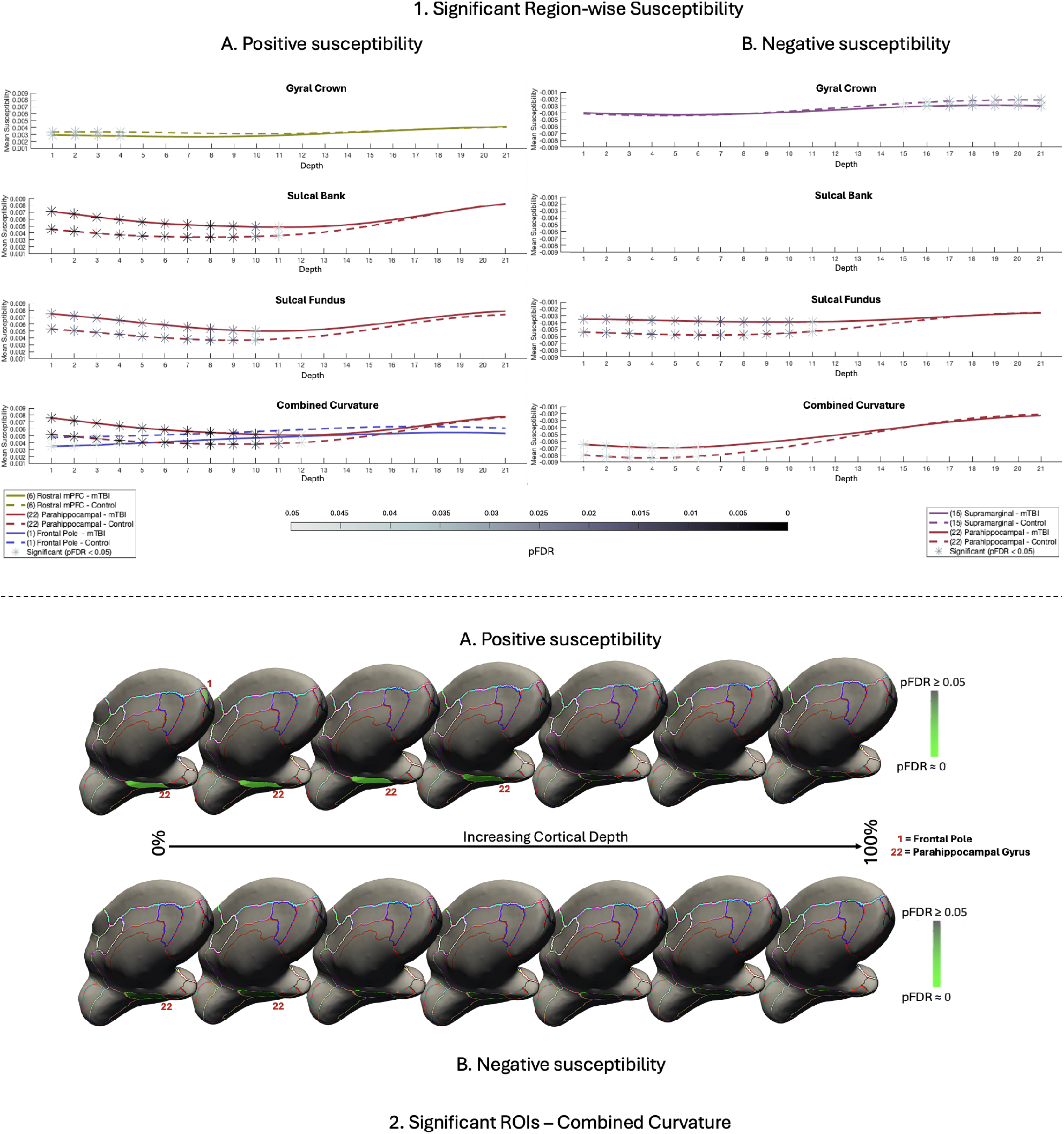
Significant region-wise susceptibility values. 1. Differences in susceptibility values between mTBI participants and controls for significant regions of interest only. A. Positive susceptibility values were significantly higher for participants with mTBI than healthy controls in superficial depths of the sulcal bank and fundus of the parahippocampal gyrus, as well as when curvature was combined. Values were significantly less positive for mTBI participants superficially in the gyral crown of the rostral mPFC and the superficial frontal pole when curvature was combined. B. Susceptibility values were significantly less negative for mTBI participants than controls in the superficial depths of sulcal fundus in the parahippocampal gyrus, as well as when curvature was combined. Values were significantly more negative after mTBI deep in the supramarginal gyral crown. Susceptibility is measured in parts per million (ppm). mPFC = middle prefrontal cortex. 2. Medial visualisation of significant ROIs after mTBI for both positive (A) and negative (B) susceptibility maps when curvature was combined as a whole-ROI measure. Surface lines demarcate borders between ROIs, projected onto an inflated surface. Significant ROIs are filled; intensity values relate directly to pFDR significance level. For visualisation purposes, depth was reduced from 21 to 7 by averaging pFDR values every 3 consecutive depths where 0% is proximal to the pial surface and 100% to the GM/WM interface.

#### Negative susceptibility

Across bilateral depth profiles, participants with sr-mTBI exhibited significantly less negative susceptibility than controls in the temporal lobe only (see *Fig. 2B*). This finding was focal to the superficial depths of the fundus in the parahippocampal gyrus, as well as when curvature was combined (see also *Fig. 3*.*1B* and *Fig. 3*.*2B*). Negative susceptibility was more negative for participants with sr-mTBI deep in the supramarginal gyral crown of the parietal cortex only. No significant differences between groups for curvature were found in the sulcal bank (see *Fig. 3B*). No significant differences were found in bilateral frontal, occipital, or insular lobes.

### Bilateral regional correlations

We used partial Pearson correlation coefficients to examine the relationship between age and regional depth-wise susceptibility (positive and negative) for combined curvature at each ROI across the entire sample, whilst controlling for group status. Additionally, we explored the correlation between susceptibility values and 1) BIST scores (Theadom et al., 2021) as an indicator of injury severity, and; 2) DSI at the time of the MRI scan as a marker of injury latency, in the mTBI sample only (see *Fig. 4*). Negative susceptibility was transformed to absolute values to better represent relationships between variables (see *Fig. 4B*). P-values were adjusted for 21 cortical depths within each ROI using FDR methods (Benjamini and Hochberg, 1995).

**Fig. 4.**
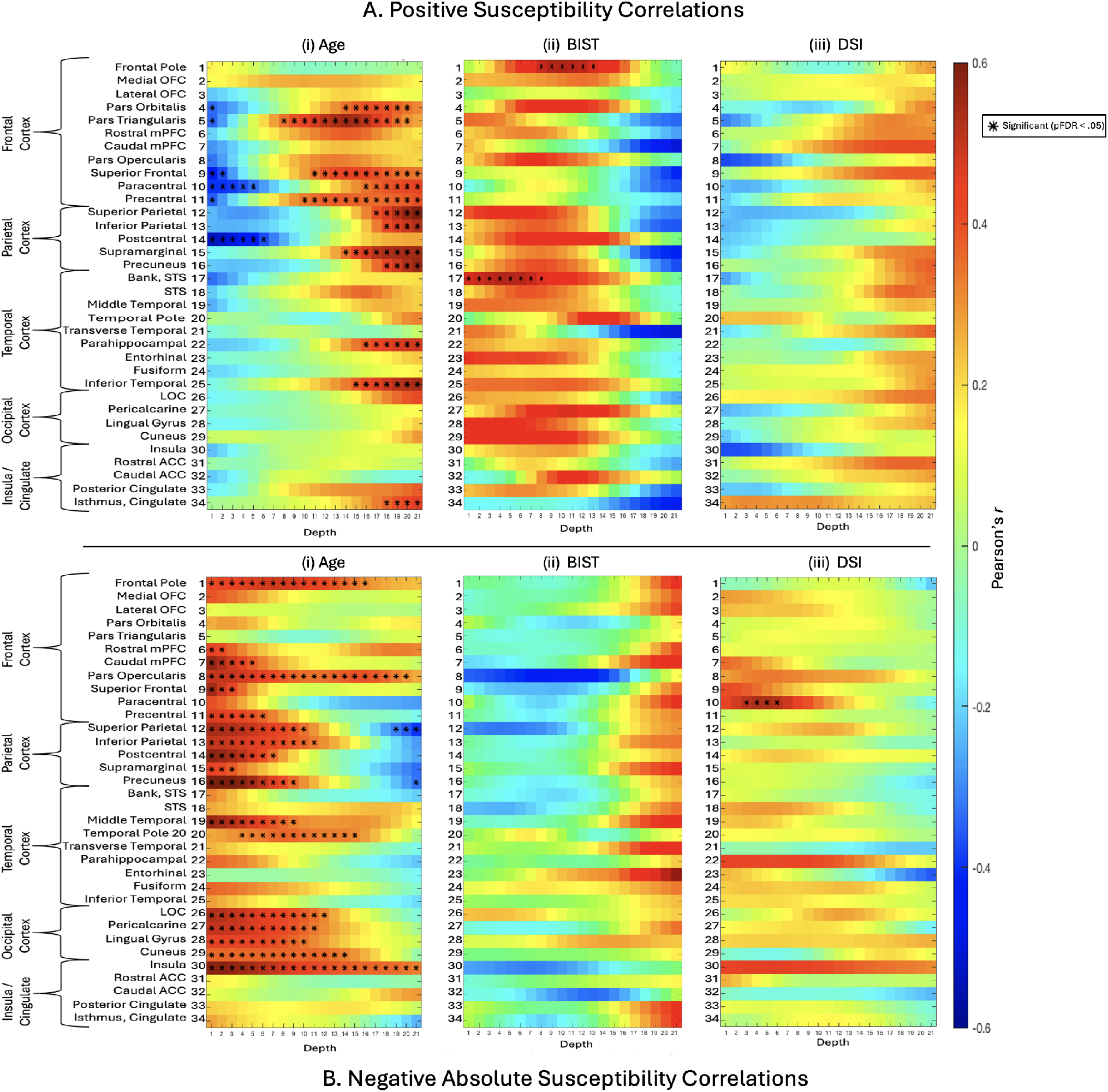
Correlations between positive or negative susceptibility and variables of interest. i: The relationship between age and positive (A) and negative (B) susceptibility values was assessed for each depth for combined curvature as a whole-ROI measure using partial correlations to control for the effects of group status. ii: the relationship between BIST scores as a marker for injury severity and susceptibility sign values was explored, along with; iii: the relationship between susceptibility sign values and DSI at the time of MRI scan as a marker of injury latency. Negative (B) susceptibility was transformed into absolute values to better represent the relationship between variables. BIST = Brain Injury Screening Tool; DSI = days since injury (at time of MRI scan).

#### Age

##### Positive susceptibility

We observed significant positive correlations between age and iron-related positive susceptibility distributions exclusively at deeper cortical depths near the GM/WM interface (see *Fig. 4A(i)*). In the frontal cortex, regions with statistically significant age-related increases in positive susceptibility values included the pars orbitalis, pars triangularis, superior frontal gyrus, paracentral lobule, and precentral gyrus. In the parietal cortex, significant regions were the superior and inferior parietal lobules, supramarginal gyrus, and precuneus. In the temporal lobe, significant positive correlations were found in the parahippocampal gyrus and inferior temporal gyrus. The only cingulate region to exhibit a positive age-related correlation was the isthmus. No significant positive correlations with age were identified in any ROIs within the occipital cortex. Conversely, we identified significant negative correlations between age and positive susceptibility exclusively at superficial cortical depths near the pial surface, demonstrating an inverse pattern to the positive correlations. In the frontal cortex, the areas showing significant negative correlations were the pars orbitalis, pars triangularis, superior frontal gyrus, paracentral lobule, and precentral gyrus. In the parietal cortex, the postcentral region exhibited significant negative correlations. No significant negative correlations between positive susceptibility and age were apparent in temporal, occipital, or insular ROIs.

##### Negative susceptibility

Significant positive relationships were observed between absolute negative susceptibility values and age, primarily in the superficial depths of ROIs across all lobes, which at times extended to the GM/WM border (see *Fig. 4B(i)*). In the frontal lobe, ROIs exhibiting significant positive correlations between age and negative susceptibility included the frontal pole, rostral mPFC, caudal mPFC, pars opercularis, superior frontal gyrus, and precentral gyrus. All examined ROIs in the parietal cortex exhibited significant positive relationships, namely, in the superior parietal lobule, inferior parietal lobule, postcentral gyrus, supramarginal gyrus, and precuneus. In the temporal cortex, significant positive correlations were found in the middle temporal gyrus and temporal pole. All occipital ROIs showed significant positive correlations, including the LOC, pericalcarine cortex, lingual gyrus, and cuneus. Within the insular cortex, only negative susceptibilities in the insula demonstrated a significant positive relationship with age. In contrast, significant negative correlations with age were observed only in the deeper cortical layers closer to the GM/WM junction of parietal regions, specifically in the superior parietal lobule and precuneus.

##### Injury severity

We observed significant positive correlations between BIST scores and positive susceptibility values in the mid-depths of the frontal pole and in superficial depths of the bank of the superior temporal sulcus (STS) (see *Fig. 4A(ii)*). No significant correlations were observed between negative susceptibility and BIST scores (see *Fig. 4B(ii)*).

##### Injury latency

No significant correlations between DSI at time of MRI and positive susceptibility values were observed for any ROI (see *Fig. 4A(iii)*). Negative susceptibility and DSI were positively correlated in superficial depths of the bilateral paracentral gyrus in the frontal cortex only (see *Fig. 4B(iii)*).

## Discussion

Previous research seeking to understand the role of brain iron following mTBI has focused primarily on susceptibility distributions in subcortical or global grey matter and/or white matter, neglecting the vulnerability of cortical regions to microstructural damage following an mTBI. The one investigation inclusive of cortical ROIs was constrained by macroscopic voxel-wise techniques, which lack the anatomical precision necessary to detect depth-or curvature-specific differences in magnetic susceptibility. To address this gap in the literature, we adapted a new analytic technique already demonstrating efficacy in Alzheimer’s disease research to perform the first investigation of sr-mTBI-related differences in magnetic susceptibility as a function of cortical depth and curvature. In line with contemporary approaches in QSM research, we separated positive (iron-related) from negative (myelin-, calcium-, and protein-related) sources of susceptibility to enable more accurate estimation of underlying biological substrates.

Our findings revealed increased positive susceptibility exclusive to the temporal lobe, specifically in the superficial depths at sulcal curvatures in the bilateral parahippocampal gyrus. This pattern was conserved when curvature was aggregated as a whole-ROI measure. In contrast, age-related positive susceptibility indicative of iron deposition was observed at deep cortical depths, closer to the interface with the white matter, in the overall sample. These findings suggest that increases in positive susceptibility close to the cortical surface indicate abnormal, injury-related iron accumulation after mTBI. Results corresponded with analyses of negative susceptibility, which demonstrated less diamagnetism in the superficial depths of the sulcal parahippocampal gyrus after mild brain trauma. In addition, negative susceptibility was positively correlated with age in superficial depths only, suggesting an alignment with age-related calcification processes in superficial cortical layers that is well supported by the literature; a pattern opposite to mTBI-related negative susceptibility effects. Fewer correlations were found between subjective injury status or time elapsed since injury and susceptibility, supporting a body of research demonstrating little relationship between objective injury measures and subjective self-report.

### Depth

*In-vitro* histological studies have demonstrated variations in iron distribution relative to specific cortical laminae (e.g., Perls iron staining), where concentrations are lowest at the pial surface and increase progressively through GM toward its junction with WM (Fukunaga et al., 2010). These findings have been corroborated by iron-sensitive R_2_* mapping of *ex-vivo* tissue samples using ultra-high field (7T) MRI (Fukunaga et al., 2010), which also exhibit high congruence with *in-vivo* QSM (Lee et al.). Taken together, these studies suggest that iron density in the cortex reflects distinct cyto- and myelo-architecture, with variance between layers. In healthy populations, iron density should be sparse at the pial surface and increase with depth. Conversely, our findings indicate an abnormal distribution pattern of positive susceptibility related to iron deposition exclusively in superficial depths of the parahippocampal gyrus in temporal cortex following injury (see *Fig. 2A, Fig. 3*.*1A*, and *Fig. 3*.*2A*). This increased positive susceptibility at the acute stage of mTBI is directly inverted for depth comparative to healthy layer-specific variation (Fukunaga et al., 2010, Lee et al.) and patterns related to normal ageing in this range of the lifespan (see *Fig. 4A(i)*), suggesting an injury-specific model of cortical microstructural trauma (see *Fig. 5*).

**Fig. 5.**
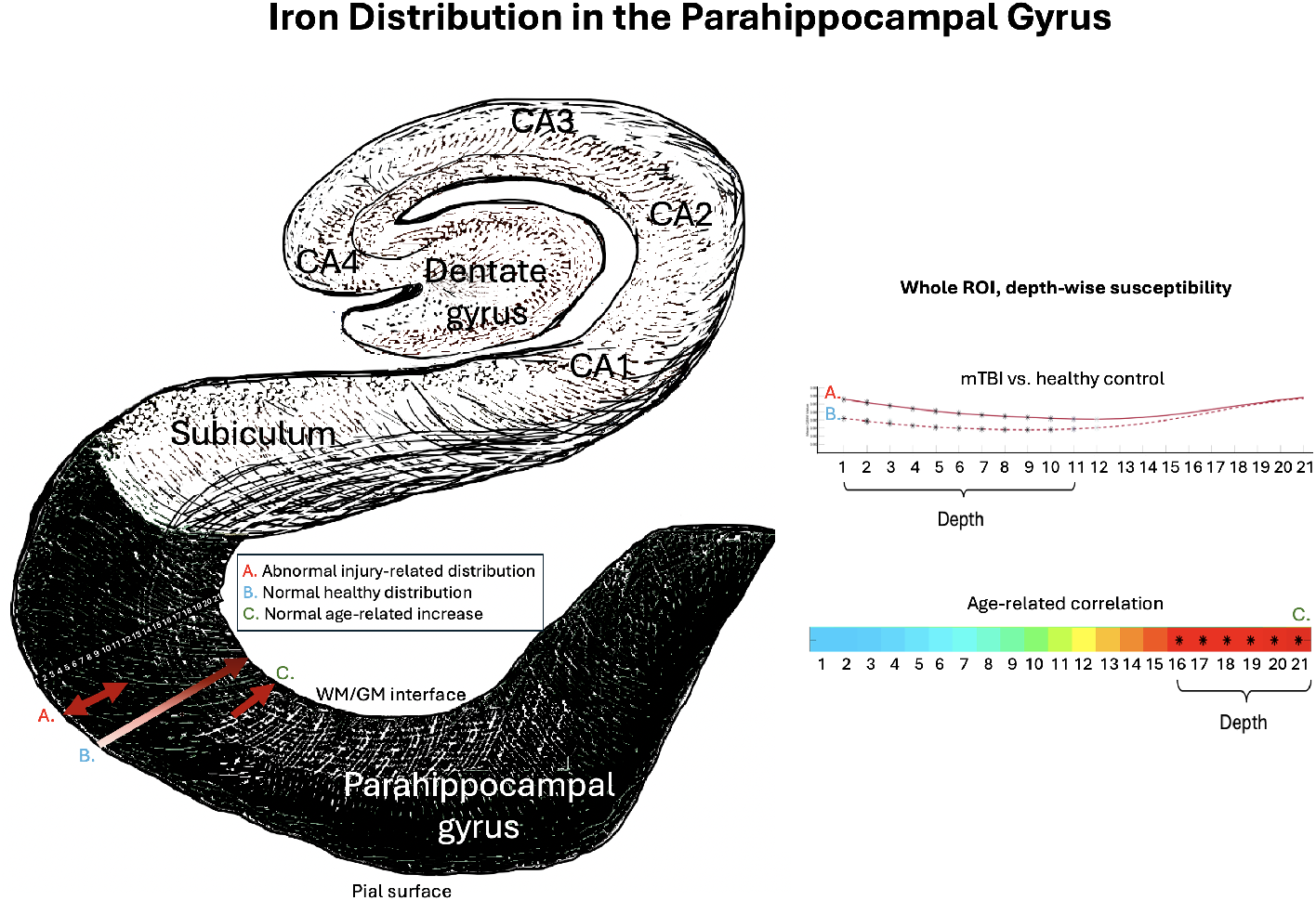
Iron and positive susceptibility distributions in the parahippocampal gyrus. Visualisation of depth-specific positive (iron-related) susceptibility distributions. (A) Depicts increased positive susceptibility following mTBI relative to healthy controls restricted to superficial depths of the parahippocampal gyrus. This pattern does not overlap with (B) validated patterns of lower iron concentration proximal to the pial surface, and increasing towards the GM/WM interface as a pattern of normal layer-specific distributions, or; (C) normal age-related increases in positive susceptibility in this population (16-32 years) restricted to the deeper cortical depths closer to the GM/WM boundary. Depiction of the hippocampal formation inspired by illustration in Ranson & Clark (1959). *The anatomy of the nervous system: Its development and function* (10th ed.). W. B. Saunders Co.

The focal nature of increased positive susceptibility in superficial depths suggests iron accumulation near small blood vessels (McKee et al., 2013). Concordantly, microhemorrhage and aggregation of activated microglia around perivascular sites are well documented after mTBI (Huang et al., 2021, Mckee and Daneshvar, 2015, Nisenbaum et al., 2014). Here, it is tempting to speculate about potential mechanisms of iron overload following mTBI. Non-heme iron (i.e., bound to proteins such as ferritin and transferrin) generally enters the brain through active transport across the blood-brain-barrier (BBB) via vascular endothelial cells; a constant process that is closely regulated within the central nervous system to ensure homeostasis (Gozt et al., 2021). Injury-induced acute cerebral microvascular dysfunction and increased permeability of the BBB (Sandsmark et al., 2019, Wu et al., 2020), likely increases iron transport into these superficial cortical layers (Levi et al., 2024, Ward et al., 2014b) which could result in a perivascular accumulation of iron. Notably, redox reactions involving non-heme iron are a significant source of reactive oxygen species (ROS) (Nisenbaum et al., 2014); when labile iron accumulates to pathological levels, it can exceed the capacity of storage proteins inducing oxidative stress, protein and DNA damage, and ferroptosis (Kruszewski, 2003, Nisenbaum et al., 2014, Ward et al., 2014b). This labile iron is frequently considered a critical contributor to secondary injury mechanisms (Gozt et al., 2021, Huang et al., 2021, Nisenbaum et al., 2014) and represents the primary source of paramagnetic susceptibility in the brain on QSM (Langkammer et al., 2012). As such, iron accumulation acting as a catalyst for auto-toxic circuits (Levi et al., 2024). is a candidate hypothesis by which iron dyshomeostasis after mTBI could account for the increased magnetic susceptibility of these compromised perivascular regions on QSM. Iron overload may represent a promising early marker of acute cell damage and mTBI-related neuropathology as well as degeneration of neural tissue downstream. However, as the precise mechanisms by which iron is released in the brain are still under investigation (Gozt et al., 2021, Zecca et al., 2004), these mechanisms cannot yet be disambiguated.

Whilst the relationship between positive susceptibility and iron is well established, the mechanisms underlying diamagnetic sources of contrast are less well understood and more challenging to elucidate (Madden and Merenstein, 2023, Merenstein et al., 2024, Northall et al., 2023). Recent research indicates that the primary source of diamagnetic contrast is myelin (Deh et al., 2018), but contributions are also made by calcium (Jang et al., 2021, Kim et al., 2020, Wang et al., 2017) and the deposition of proteins such as amyloid-beta (A*β*) (Gong et al., 2019, Zhao et al., 2021). Our results suggest that negative susceptibility is decreased in the parahippocampal gyrus at superficial depths following mTBI. Myeloarchitecture studies indicate that Layer I of cerebral cortex consists primarily of axons, dendrites, and axon terminals, the cell bodies of which are located in deeper layers (Miyashita, 2022). Although the cerebral cortex is not well known for high myelin content (Northall et al., 2023), decreased positive susceptibility at these superficial depths could indicate changes to surface axons. This pattern coincided with increased positive susceptibility in the superficial parahippocampal gyrus, suggesting a dual model of perivascular microstructural trauma focal to this region. The majority of iron present in the human brain parenchyma is stored as non-heme iron within myelin-maintaining oligodendroglia and in myelin itself (Connor and Menzies, 1995; pathological iron levels have been known to damage both (Bradl and Lassmann, 2010, Hametner et al., 2013, Lassmann et al., 2012). Mechanisms leading to co-localisation of increased positive susceptibility and decreased negative susceptibility observed in this study are further supported by research into pathogenesis of multiple sclerosis, which indicates that neuroinflammation and altered BBB permeability can result in iron accumulation in macrophages and other iron-related cytotoxic events, such as oxidative stress, causing degradation of oligodendrocytes and axons (Bradl and Lassmann, 2010, Lassmann et al., 2012, Ward et al., 2014b). Conversely, increased negative susceptibility was mostly observed at deeper cortical depths closer to the GM/WM junction, which might reflect aggregation of A*β* inherent to AD (Braak and Braak, 1991), and sometimes present in CTE (McKee et al., 2023), or calcifications known to negatively affect cognition (Thibault et al., 2007). Notably, the injury-related accumulation patterns did not overlap with age-related changes in negative susceptibility, which increased at superficial depths (see *Fig. 4B(i)*). This suggests that both decreased negative susceptibility in superficial depths and increased negative susceptibility at deeper cortical depths may result from abnormal, injury-related, neuropathological processes.

### Curvature and ROI

Prior research has noted that the sulcal fundus is particularly vulnerable to injury in mTBI due to increased susceptibility to mechanical deformation (McKee et al., 2023, Smith et al., 2013) and the ‘water hammer effect’(Kornguth et al., 2017). The current study supports a fundus-specific model of damage in mTBI; the only persistent differences between groups in both positive and negative susceptibility were observed in sulcal regions of the bilateral parahippocampal gyrus (positive susceptibility: sulcal bank and fundus; negative susceptibility: sulcal fundus (see *Fig. 2* and *Fig. 3)*). The focal nature of injury to these concave regions extends prior work noting increased mean cortical curvature in the sulcus (King et al., 2016) and sulcal widening (Kornguth et al., 2017) after mTBI. Results are also consistent with observations of mTBI-related vascular injury and microhemorrhage in the sulcal fundus on SWI (Kornguth et al., 2017). Additionally, the medial temporal lobe is a notable site of injury in sr-mTBI and previous research has highlighted the associations between sr-mTBI and loss of cortical thickness in the parahippocampal gyrus along with reductions in parahippocampal volume (Arciniega et al., 2024). Contextually, the parahippocampal cortex acts as a hub region in a network connecting areas of the frontal, parietal, and temporal lobes (Raslau et al., 2015) and represents a vital link between the default-mode network and the medial temporal lobe memory system, as evidenced by resting-state fMRI studies (Ward et al., 2014a). As such, it is integral to various cognitive processes including visuospatial processing and episodic memory (Aminoff et al., 2013) and the facilitation of contextual associations fundamental to higher order cognitive performance (Raslau et al., 2015). The co-localisation of iron-related deposition and potential myelin changes, particularly in integrative superficial regions, within the temporal parahippocampal gyrus is consistent with the memory impairments symptomatic of sr-mTBI (Mckee and Daneshvar, 2015).

### Age and clinical correlates

Increasing iron in deep grey matter nuclei and some regions of the cortex is a hallmark of normal ageing (Zecca et al., 2004, Hallgren and Sourander, 1958). Research indicates that age-related iron increases occur primarily in the motor and premotor cortices, as well as the superior prefrontal and parietal cortices (Acosta-Cabronero et al., 2016, Hallgren and Sourander, 1958). Additionally, these increases have been reported in the insula (Acosta-Cabronero et al., 2016) and hippocampus (Hagemeier et al., 2012). Histological evidence suggests that normal increases in cortical non-heme iron may be especially pronounced in younger individuals, sharply increasing during childhood and plateauing at around 30 years of age, depending on region (Hallgren and Sourander, 1958, Schenck and Zimmerman, 2004). These observations are supported by some (Acosta-Cabronero et al., 2016, Callaghan et al., 2014) but not other (Rodrigue et al., 2011) crosssectional iron-sensitive MRI findings in adjacent age groups. Our findings support age-related cortical iron increases in a youthful population. In line with prior research, ROIs with age-related increases in likely iron content were most dense in frontal and parietal lobes, in addition to two loci in the temporal lobe and one in the insular cortex (see *Fig. 4A(i)*). Of all cortical regions, the motor system is particularly affected by age-related iron deposition (Acosta-Cabronero et al., 2016), a pattern mirrored in our findings which show a notable density in the primary motor cortex. Our results suggest that age-related increases are restricted to deeper cortical depths closer to the GM/WM junction, which are known to naturally express higher iron levels in the healthy population (Fukunaga et al., 2010, Lee et al.). These deeper layers contain large pyramidal cells (Miyashita, 2022), which could speculatively accumulate iron differently. Indeed, previous research (Merenstein et al., 2024) has suggested that because age is more predictive of delayed response time rather than difficulties in decision-making (Ratcliff, 2008, Madden et al., 2020, Merenstein et al., 2023), age-related iron accumulation should show a preference for deeper layers responsible for output, rather than more integrative superficial regions (Rolls and Mills, 2017). In addition, we observed age-related increases in negative magnetic susceptibility exclusively close to the pial surface. Previous studies utilising negative QSM have shown similar distributions at superficial depths (Northall et al., 2023). This pattern is well-supported in the literature, as research indicates that calcifications commonly occur in areas with high vascularization and metabolic activity (Jang et al., 2021), such as the cortical surface. Taken together, our findings indicate that age-related increases in iron are likely restricted to deep cortical depths, while diamagnetic calcifications are likely to occur at superficial depths. Both observations are supported by strong biological plausibility and well-documented scientific evidence. This reinforces the conclusion that observed differences in both positive and negative susceptibility following sr-mTBI reflect pathological processes distinct from those associated with normal ageing.

The general lack of correlation between positive or negative susceptibilities and BIST scores may be attributed to the under-reporting of symptoms notorious in sr-mTBI (Meier et al., 2015), which would preclude accurate correlations. Whilst iron negatively correlates with cognitive symptoms in mTBI (Lu et al., 2015), it is well-documented that subjective assessments of injury severity do not reliably reflect objective brain injury or recovery status (McCrea et al., 2017) and cognitive/clinical symptoms do not align with neuroimaging findings (Shenton et al., 2012). Additionally, the lack of correlation between positive or negative susceptibilities and injury latency suggests that acute neuropathology may persist into the subacute and chronic stages, necessitating further research to elucidate the time course of secondary injury in sr-mTBI.

### Implications

The role of iron in acute responses following cytotrauma, as well as its co-localisation with, and involvement in, misfolded proteins in neurodegenerative diseases, including CTE, identifies it as a potential marker of early degenerative processes (Zecca et al., 2004, Bouras et al., 1997). Interestingly, the temporal lobe and hippocampus are primary loci of atrophy and tau deposition in CTE (Murray et al., 2022, McKee et al., 2023), and iron has been found in neurofibrillary tangles within these regions (Bouras et al., 1997). In CTE, these neurofibrillary tangles favour superficial cortical layers (II/III) (Pearce et al., 2020, Tokuda et al., 1991) and aggregate around small blood vessels (McKee et al., 2016), especially at the sulcal fundus (Smith et al., 2013). This laminar predilection is not seen in other tauopathies, like AD, where tau phosphorylation generally occurs in deeper layers (V/VI) (Pearson et al., 1985). Indeed, this specific distribution is considered pathognomonic of CTE (McKee et al., 2016, 2023). Notwithstanding the short-term effects of iron overload, which have been linked to secondary injury in mTBI (Huang et al., 2021, Nisenbaum et al., 2014), the association between iron-mediated oxidative stress and hyperphosphorylation of tau (Yamamoto et al., 2002) coupled with the similarity of iron distributions observed in this study to hallmark distributions of tauopathy and degeneration in CTE raises important questions about the disease path from acute injury to eventual tissue degeneration. The precise mechanisms underlying CTE remain an active area of research, but these findings suggest that iron may serve as a candidate biomarker, warranting further investigation. Until such a time, any parallels drawn between the pattern of iron distribution in this study and the hallmark features of CTE tauopathy can, at this stage, only be speculative.

Here, follow-up investigations will be of paramount importance. A better understanding of iron and other neurobiological correlates, such as myelin damage or protein aggregation, may be essential for developing effective care and targeted treatments at the acute stage of mTBI, which could protect against adverse consequences later in life. Recent research has explored the possibility of using heavy metal chelation as a therapeutic target for iron in AD (Mazur et al., 2024) and TBI (Daglas and Adlard, 2018). In AD, this approach has had limited success, perhaps due to the accumulation of iron over a long time course; once iron overload becomes apparent, cell death has already occurred (Levi et al., 2024). However, mechanisms of acute injury-related iron overload may be distinct from long-term aggregation. Murine models using iron chelators such as Deferoxamine (Jia et al., 2023, Long et al., 1996, Panter et al., 1992, Zhao et al., 2014) and *N,N* -Di(2-hydroxybenzyl)ethylenediamine-*N,N* -diacetic acid monohydrochloride (HBED) (Khalaf et al., 2019) have shown promise in reducing TBI symptomatology, likely through inhibition of ferroptosis and reductions in neuroinflammation, ROS, and gliosis. More research is needed to understand efficacy in humans, but this remains a possible avenue for limiting the effects of acute iron dyshomeostasis which may contribute to degenerative effects evident in later life.

### Limitations and future research

While QSM offers valuable insights into iron content and distribution, at lower field strengths (i.e., 3T) voxel resolution is restricted and does not reflect specific architectonics of cortical tissue. Cortical column analyses were thus constrained to investigations of depth rather than susceptibility distribution specific to cortical laminae. Future research could benefit from applying cortical column analysis to images with higher resolutions, for example those collected on high field scanners, or by pushing images to sub-1mm resolutions insofar as signal-to-noise ratios are not compromised. The present study utilised a single-echo QSM sequence, limiting thresholding of susceptibility sources to *between* voxels. To enable thresholding *within* voxels, future studies should incorporate multi-echo sequences (Lee et al., 2023, Li et al., 2023) where available, thereby enhancing the biological interpretation of the findings. Incorporating complementary modalities, such as positron emission tomography (PET), would provide a more comprehensive understanding of cellular metabolism and neurobiological processes, potentially linking these to the observed mTBI-related differences and enabling more biologically informed inferences. Inclusion of protein assays, such as blood-based biomarkers, would also aid in creating a more comprehensive picture of the biological consequences of sr-mTBI. A range of potential confounds were not controlled for in this study including prior injuries, genetic predispositions, and environmental influences, which may affect injury severity and presentation (Rosenbaum and Lipton, 2012), and future research should consider including additional measures to control for these variables. Given that this study was conducted exclusively in a male cohort, the generalisability of the findings to females may be limited. Sex differences in injury effects are reportedly influenced by a range of factors, including hormonal variations (Wunderle et al., 2014), the use of oral contraceptives (Gallagher et al., 2018), and differences in neck musculature (Tierney et al., 2005). Future research should extend investigations to female athletes and consider comparisons by sex. Finally, given the limitations of inferring long-term consequences of sr-mTBI from data collected at the acute stage, future research should prioritise longitudinal studies tracking athletes over time. This prospective approach would yield more precise information compared to retrospective studies, providing valuable insights into the relationships between the number of mTBIs sustained, iron accumulation, recovery trajectories, and long-term outcomes. In lieu of longitudinal research, planned upcoming studies will utilise comparison of individual z-scores relative to healthy population norms to better clarify the prevalence of adverse outcomes following sr-mTBI. This approach will aim to enhance our understanding of the impact of sr-mTBI at the individual level, which may be obscured by group-level analyses (Bedggood et al., 2024, Domínguez D et al., 2024).

## Conclusions

To better characterise the mechanisms of mild traumatic brain injury at the acute stage, we conducted the first QSM study to assess depth- and curvature-specific regional patterns of positive (iron-related) and negative (myelin-, calcium-, and protein-related) magnetic susceptibility in the cerebral cortex following injury. We observed co-localisation of increased positive susceptibility with decreased negative susceptibility in the parahippocampal gyrus, indicating a possible accumulation of iron concomitant with myelin changes after injury. This distribution pattern was observed in the superficial depths of the sulcus, suggesting perivascular trauma due to mechanical forces. The pattern appeared distinct from age-related differences in positive and negative susceptibility and was reminiscent of pathognomonic patterns of tau pathology in CTE, which is known to co-localise with iron. These results support a complex, dual-pathology model of trauma after mild head injury and have implications for understanding microstructural brain tissue damage following mTBI.

## Author Contributions Statement

**Christi A. Essex** (Conceptualization, Methodology, Project Administration, Validation, Software, Formal Analysis, Investigation, Resources, Data Curation, Writing - Original Draft, Writing - Review & Editing, Visualization); **Jenna L. Merenstein** (Methodology, Writing - Review & Editing, Visualization); **Devon K. Overson** (Methodology, Software, Visualization, Validation, Writing - Review & Editing); **Trong-Kha Truong** (Methodology, Software, Writing - Review & Editing); **David J. Madden** (Methodology, Software, Writing - Review & Editing); **Mayan J. Bedggood** (Writing Review & Editing, Project administration, Investigation); **Helen Murray** (Writing - Review & Editing); **Samantha J. Holdsworth** (Writing - Review & Editing); **Ashley W. Stewart** (Writing - Review & Editing); **Catherine Morgan** (Writing - Review & Editing); **Richard L.M. Faull** (Writing Review & Editing); **Patria Hume** (Writing - Review & Editing); **Alice Theadom** (Conceptualization, Methodology, Writing - Review & Editing, Funding acquisition, Supervision); **Mangor Pedersen** (Conceptualization, Methodology, Writing Review & Editing, Funding acquisition, Supervision)

## Acknowledgments

We extend thanks to Aria Courtney, Amabelle Voice-Powell, and Cassandra McGregor for their contribution to the data collection, and Tania Ka’ai for bringing her perspective to cultural considerations on this study. In addition, we thank Axis Sports Concussion Clinics, particularly Dr Stephen Kara, for their assistance with recruiting sr-mTBI participants and personnel at the Centre for Advanced Magnetic Resonance Imaging (CAMRI) for their assistance collecting MRI data. We also acknowledge Dr Tim Elliot for radiological reporting of all participants and Siemens Healthcare/Siemens Healthineers for the use of a work-in-progress (WIP) sequence for the acquisition of magnitude and phase images.

## Data availability

De-identified MRI data and code used for image processing and statistical analysis can be made available upon request to the corresponding author. Parent codes for cortical column generation can be made available upon request from co-authors (JLM, TKT) based at the Brain Imaging and Analysis Center at Duke University Medical Center.

